# Development and validation of AI-based pre-screening of large bowel biopsies

**DOI:** 10.1101/2022.11.30.22282859

**Authors:** Mohsin Bilal, Yee Wah Tsang, Mahmoud Ali, Simon Graham, Emily Hero, Noorul Wahab, Katherine Dodd, Harvir Sahota, Shaobin Wu, Wenqi Lu, Mostafa Jahanifar, Andrew Robinson, Ayesha Azam, Ksenija Benes, Mohammed Nimir, Katherine Hewitt, Abhir Bhalerao, Hesham Eldaly, Shan E Ahmed Raza, Kishore Gopalakrishnan, Fayyaz Minhas, David Snead, Nasir Rajpoot

## Abstract

**Background:** Histopathological examination is a pivotal step in the diagnosis and treatment planning of many major diseases. With the aims of facilitating diagnostic decision-making and improving the use of pathologists’ time, we developed an AI-based pre-screening tool that analyses whole slide images (WSIs) of large bowel biopsies to identify normal, inflammatory, and neoplastic biopsies.

**Methods:** To learn the differential histological patterns from digitised WSIs of large bowel biopsy slides stained with Haematoxylin and Eosin (H&E), our proposed weakly supervised deep learning method uses only slide-level diagnostic labels and no detailed cell or region-level annotations. The proposed method was developed on an internal cohort of biopsy slides (n=5054) from a single laboratory labelled with corresponding diagnostic categories assigned by pathologists. Performance of the tool was evaluated on the internal development cohort (n=5054) in a cross-validation setting, and three external unseen cohorts (n=1536) for independent validation.

**Findings:** The proposed tool demonstrates high degree of accuracy to assist with the pre-screening of large bowel biopsies, being able to identify neoplastic biopsies (AUROC = 0·993), inflammatory biopsies (AUROC = 0·966) and all abnormal biopsies (AUROC = 0·979). On the three independent validation cohorts, it achieves AUROC values of 0·943, 0·958 and 0·964 for the detection of abnormal biopsies. Analysis of saliency maps confirms the representation of disease heterogeneity in model predictions and their association with relevant histological features. Interestingly, after examining diagnostic discrepancies between the proposed AI tool and original diagnostic labels, a panel of pathologists found that the proposed tool correctly identified a number of abnormal slides that had been initially reported as normal.

**Interpretations:** The proposed tool with its high sensitivity of detecting abnormal colorectal biopsies promises significant improvements in clinical workflow efficiency and assistance in diagnostic decision-making through pre-screening of normal biopsies.

**Funding:** Innovate UK on behalf of UK Research and Innovation.

## Introduction

Colorectal cancer (CRC) is the fourth most common cancer in the UK and the 2^nd^ leading cause of cancer-related fatalities in the UK and US (1,2). Over 42 000 people are diagnosed with CRC every year in the UK^2^ and 140 000 in the US. Most CRCs develop from polyps, a pre-cancerous outgrowth of tissue from the lining of the colon. Colonoscopy has long been the reference standard for investigation to examine the entire rectum and colon for precancerous polyps, tumours or other problems (3,4). A tissue sample (biopsy) taken during colonoscopy helps make a definitive diagnosis of any colonic abnormalities. Microscopic examination of histopathological features of colorectal biopsies is the ‘gold standard’ to provide to the clinicians with a definitive diagnosis of any colonic pathology that may be found. A wide range of colonic abnormalities can be found, including carcinoma, polyps (non-neoplastic or neoplastic), inflammation and ulceration caused by infection, medications, inflammatory bowel disease and microscopic colitis.

Improvements in and increased roll-out of large bowel cancer screening programs, designed to detect CRC at the earliest stage, have contributed to an increase in the pathologist workload. Besides, early-stage disease is often more difficult to detect than late-stage disease. In addition, the requirement for more precise diagnostic information needed to provide the best standard of care is contributing to the increased workload experienced by pathology laboratories world-wide (5). Digitisation of cellular pathology laboratories and an accelerated transition to digital pathology during the COVID-19 pandemic (6) offer an opportunity for automated pre-screening of large bowel cancer to address the pathologist shortage and improve clinical management.

Approximately 30-40% of large bowel endoscopic biopsies are reported as normal and contain no discernible pathology, resulting in pathologists spending a significant amount of their time looking for pathology that is non-existent (7). Pre-screening of colon biopsies with an Artificial intelligence (AI) tool will save pathologist time allowing the pathologists to dedicate more time on abnormal biopsies, where their expertise is needed most. This approach is quite different to the existing AI in pathology literature where screening is modelled to differentiate normal and cancer cases (8–11) or cancer and inflammatory cases (12,13) only. Current AI approaches may have benefits in improving speed of reporting, but do not remove the large volume of normal biopsies which still need to be reported by a human pathologist. AI based diagnostic tools for large bowel biopsy pre-screening to improve pathology workflow is an unmet need of high clinical relevance. We argue that developing high sensitivity AI models will lead to an effective, reliable, and low-risk AI-based pre-screening tool suitable for clinical workflow. To the best of our knowledge, there is no existing AI tool addressing pre-screening of colorectal biopsies for both non-neoplastic and neoplastic abnormalities.

In recent years, an increasing number of weakly supervised deep learning methods for whole-slide image classification have been proposed for various histopathology problems (14–20). An attractive feature of weakly supervised methods is their ability to enable automatic classification without the need for detailed pixel or region level annotations. These methods can be made to work efficiently on thousands of WSIs, often after dividing them into smaller parts as image tiles (or patches), resulting in millions of image tiles. Recently, Oliveira *et al*. (21) have identified limitations of existing algorithms and underscored the need for more accurate methodologies for use in clinical practice. They highlighted the need for larger datasets and the use of appropriate learning methodology to improve prediction accuracy. In our previous work, we proposed an iterative draw and rank sampling based weakly supervised WSI classification pipeline (IDaRS) for prediction of molecular pathways including microsatellite instability and genetic mutations of BRAF, KRAS, and TP53 in CRC (22).

#### Research in context

##### Evidence before this study

We queried Google Scholar without language restrictions and with the search terms “screening or pre-screening of (colorectal or colon or rectal) cancer or biopsies AND (machine OR deep) learning OR (artificial intelligence OR AI)” and analysed the top 50 scientific articles published between Jan 1, 2018, and Aug 30, 2022, to examine evidence before this study. Several studies have shown the use of deep learning or machine learning to identify colorectal cancer or polyps from normal slides or inflammatory slides from cancer slides directly from routine whole-slide images of tissue sections. However, previous studies did not attempt the clinical application of pre-screening colonic biopsies with high sensitivity in differentiating the two major types of abnormal colonic biopsy slides (inflammatory and neoplastic) from normal slides. Studies have also suggested that previous weakly supervised approaches did not achieve the level of accuracy needed for clinical practice.

##### Added value of this study

Motivated by an unmet clinical need, we developed a weakly supervised deep learning model for multi-class whole-slide image classification, together with confidence prediction and using only slide-level labels, for pre-screening of colon biopsies. Our method outperformed recent state-of-the-art weakly supervised classification methods for the identification of normal and abnormal colonic biopsies in both internal and external validation cohorts, yielding 56% specificity at 99% sensitivity.

##### Implications of all the available evidence

With 99% sensitivity in predicting abnormal categories, our AI tool can potentially be used in clinical workflows to automatically report normal colonoscopies promising reduction of more than half normal colon biopsies in the pathologist workload and improvements in diagnostic efficiency and patient management. After large-scale validation and enhanced domain generalisation with data from multiple cohorts, it can be implemented in multiple centres. Prediction of slide labels together with a spatial

In this study, we present a customised deep learning algorithm for pre-screening of colorectal biopsies based on digitised whole slide images (WSIs) of biopsy slides to distinguish between normal and abnormal biopsies. We propose a multi-class weakly supervised learning algorithm to distinguish between normal, inflammatory and neoplastic colonic biopsies and to learn confidence in tile-level predictions. We show that the proposed **C**olorectal **AI M**odel for **A**bnormality detectio**N** (CAIMAN) algorithm can be used for pre-screening of colon biopsies and demonstrate its high accuracy through cross-validation in a large internal cohort and external validation on entirely unseen independent cohorts. CAIMAN offers pre-screening of normal colonic biopsies, with the ability to predict the confidence value together with prediction of the slide label and to highlight regions containing neoplastic and non-neoplastic abnormalities to aid the pathologist diagnosis.

## Methods

### Study Design

This study was undertaken using an internal development cohort and three independent validation cohorts. The development cohort of large bowel biopsies was scanned at the University Hospitals Coventry and Warwickshire (UHCW) NHS trust, a large academic teaching hospital in the UK covering a local population base of over 300 000 people. We posed colorectal biopsy pre-screening as a three-class classification problem where each WSI is classified into one of the following three categories: normal, abnormal non-neoplastic and abnormal neoplastic. As shown in Figure 1, the proposed CAIMAN model was developed by training a deep convolutional neural network (CNN) for three-class classification on image tiles from training slides of the UHCW cohort. Prediction scores of image tiles were then aggregated into three prediction scores for the whole slide, one each for its likelihood of being normal, non-neoplastic, and neoplastic. The CAIMAN validation followed mostly the same steps but with a frozen CAIMAN model.

**Figure 1.**
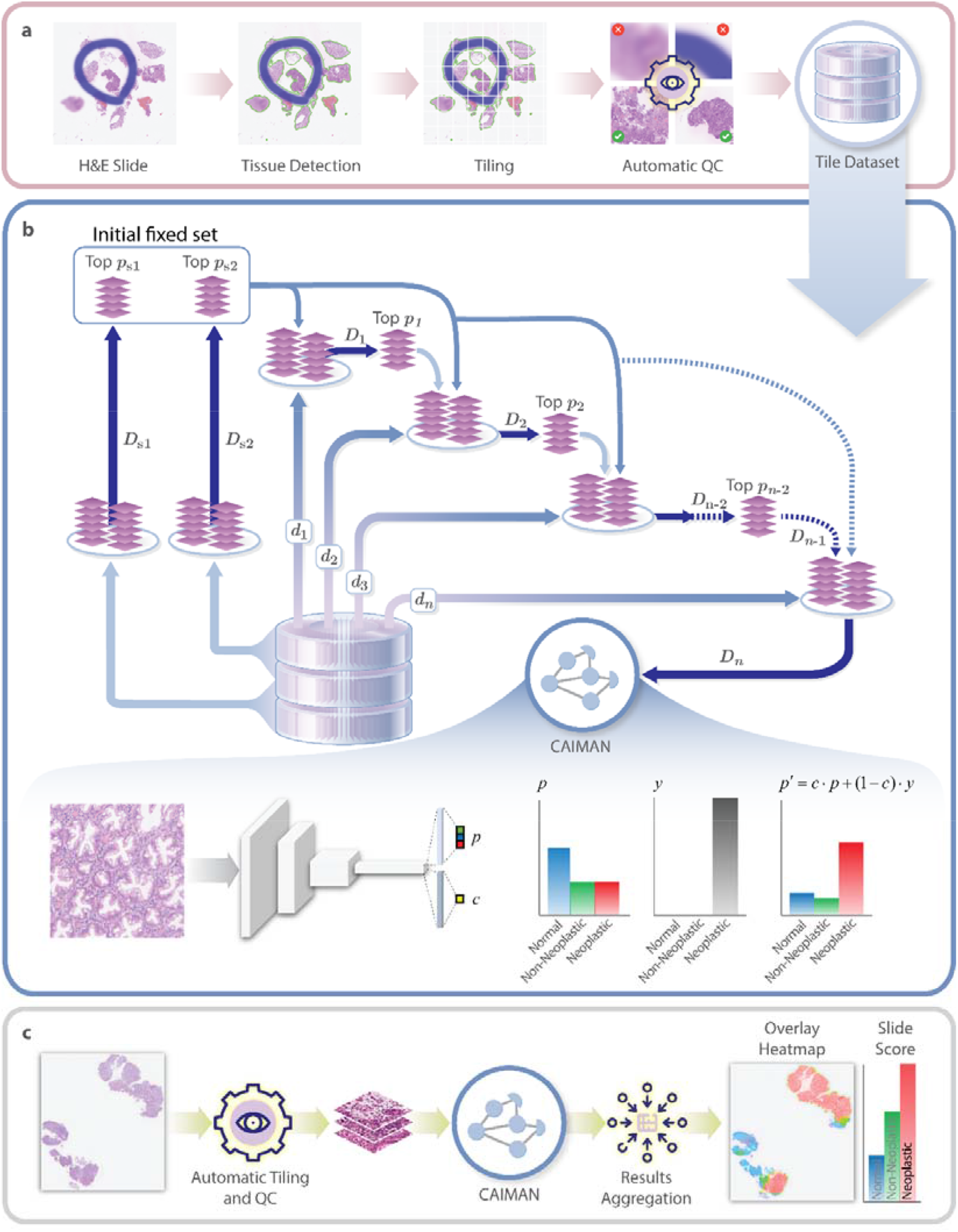
*AI based colon biopsy pre-screening using CAIMAN: a. Pre-processing. b. CAIMAN for training data per iteration c. CAIMAN inference. D*_*i*_ : *CAIMAN* deep learning model at *i*^*th*^ training iteration. *d*_*i*_: Draw subset of image tiles. Top *p*_*i*_: Top 50% tiles at *i*^*th*^ training iteration, where, *i = 1,2,3,…,n. p*: tile’s probability of belonging to diagnostic categories produced by *CAIMAN. c:* tile’s confidence on its prediction produced by *CAIMAN. p′*: confidence weighted probability.

### Data collection and preparation

#### UHCW – The development and internal validation cohort

During 2019, the cellular pathology laboratory at UHCW processed a total of 41 771 histopathology requests, including 4877 (11·7%) large bowel biopsies of which 1680 (34·4%) were normal. The UHCW cellular pathology laboratory achieved the milestone of 100% digital scanning of colorectal cases in 2016 with all gastrointestinal pathologists fully validated for digital pathology reporting, except for bowel cancer screening cases, which are required to be reported using conventional glass slide microscopy (Public Health England letter of instruction). A total of 13 pathologists were involved in the development of the pre-screening tool. In developing a clinical-grade AI-based pre-screening tool, pathologists must be actively involved, and the target population should be realistically represented in the development cohort.

All slides in the UHCW cohort were diagnostic standard Haematoxylin and Eosin (H&E) stained Formalin-Fixed Paraffin-Embedded (FFPE) histology slides scanned at 40× (0·275 microns per pixels or MPP) using the GE Omnyx JP2 scanner at UHCW. The slides were reviewed by two pathologists, with additional pathologists providing consensus diagnoses when there was a discrepancy on slide-level diagnoses (normal, abnormal non-neoplastic, and abnormal neoplastic).

The data collected included the biopsy site in the colon as well as the slide-level diagnosis. Supplementary Figure S1a (Appendix p.2) shows the specimen sites and percentages of biopsies for UHCW that include transverse, ascending, descending, and sigmoid colon, caecum, and rectum. Figure S1b (Appendix p.2) lists the various types of abnormal sub-conditions present in colorectal biopsies from the broader non-neoplastic and neoplastic categories in our development and validation cohorts. The neoplastic category combines early pre-cancerous and cancerous abnormalities while the non-neoplastic category combines all other abnormal conditions (mostly inflammatory conditions) that are not neoplastic in nature.

#### Independent validation cohorts

Three external cohorts were used for independent validation of the proposed CAIMAN tool for pre-screening of colon tissue slides. External validation of the trained CAIMAN model was performed using two external cohorts from the UK which contained WSIs from East Suffolk and North Essex (ESNE) NHS Foundation Trust and South Warwickshire NHS Foundation Trust, and a third cohort of digitised WSIs with slide labels (again, without any detailed annotations) from IMP Diagnostics Laboratory, Portugal (21). Figure S2 (Appendix p. 3) shows the data selection for both the (internal) development and independent (external) cohorts.

### Data pre-processing

As shown in Figure 1a, tissue regions were segmented from each WSI to extract tiles corresponding to the tissue areas only. Square tiles of 256×256 pixels are extracted with a stride of 128 pixels (50% overlap) from a downscaled version of WSI at 5× objective magnification (corresponding to 2·2 micron per pixel or MPP) from the segmented tissue regions. A tile is kept for subsequent processing if it contains > 50% tissue specimen. WSIs with fewer than four tiles are excluded. We perform an image quality check to filter out blurry artifacts and pen marks using Canny edge detection in Python’s OpenCV package (19).

### Weakly supervised deep learning using CAIMAN

As training labels are available only at the WSI level and not all patches in a WSI are predictive of its diagnostic label, the CAIMAN model employed a weakly supervised classification pipeline building on our previously published Iterative Draw and Rank Sampling (IDaRS) algorithm (22). Like IDaRS, CAIMAN works by performing iterative draw and rank sampling of patches from a set of training WSIs. However, unlike IDaRS, CAIMAN addresses colon biopsy pre-screening as a multi-class weakly supervised learning problem and also learns to predict confidence scores.

As shown in Figure 1b, instead of using all tiles, we chose three subsets of image tiles from each slide for training. For the first two iterations, we split the whole training set into two sets by randomly dividing tiles of each slide into two halves, one for the first training iteration and another for the second iteration. We obtain the top 50% tiles from the first two iterations and combine them into an initial fixed set for rest of the training iterations. For the next and subsequent training iterations, we have an initial fixed subset of top tiles, a subset of randomly drawn proportion (d) of tiles and a subset of top-ranked proportion (*k*) of the training tiles. The parameters for CAIMAN were empirically chosen as follows: *d=50%* and *k=50%*, maximum of 10 training iterations and batch size of 1024 tiles. We fine-tune the backbone ResNet34 (23) network (originally pretrained on ImageNet) on the UHCW cohort using cross entropy loss, the Adam optimizer with initial learning rate and weight decay of 10^−4^.

For weakly supervised training in an end-to-end manner, we added another output branch of a single neuron in the backbone network that learns to estimate confidence for each tile’s prediction along the lines of (24) as shown in Figure 1b. We modified the batch training loss by combining the tile and confidence losses with the slide loss (for abnormal slides only). Slide loss for each abnormal slide was calculated separately by averaging losses of all tiles of the abnormal slide and all normal tiles. A mathematical description of loss function can be found in Supplementary methods (Appendix p.3). During the training process, we iteratively use CAIMAN to produce confidence weighted prediction scores for each tile and select the top-ranking *k* tiles of each slide, i.e., those with high likelihood of being abnormal.

The CAIMAN model assigned three probability values and a confidence score to each tile at the inference time (Figure2c). The probabilities correspond to the likelihood of a tile belonging to normal, abnormal non-neoplastic and abnormal neoplastic histology in a multi-class classification fashion. At inference time, we obtained confidence weighted prediction scores by multiplying confidence prediction with each of three probability values, separately. Confidence weighted scores of all tiles in a WSI were considered for average aggregation into a single WSI score, which represent inference scores of the WSI being normal, abnormal non-neoplastic and neoplastic. We then map the score of each tile to blue, green and red colours for generating overlay heatmaps, illustrating the likelihood of tile belonging to normal, non-neoplastic, and neoplastic, respectively.

We used PyTorch for implementation of our CAIMAN deep learning model. A set of data augmentations – including random rotation with angles at 0, 90, 180, and 270 degrees, random horizontal and vertical flip transformations, colour jitter with brightness, contrast, saturation of 0·3 and hue of 0·05, randomly adjusted sharpness with factor 2, and random auto contrast – were applied on-the-fly on all training tiles (of size 256×256 pixels at ≈2·2 MPP). All experiments were conducted on an Nvidia DGX-2 Deep Learning System with 16×32GB Tesla V100 Volta GPUs in a shared environment. The deep learning model was built on 2 parallel GPUs with 10 worker threads, with each GPU having a dedicated RAM of 32 GB. Supplementary methods (p.3,4) provide the pseudocode of CAIMAN algorithm from a theoretical point of view.

### Statistical analysis

We performed three-fold *internal* cross-validation with case-controlled stratification for performance evaluation of CAIMAN using the internal UHCW cohort. In this evaluation protocol, the dataset was divided into three subsets based on the case identifiers to ensure that all WSIs of a case are in the same fold. For each fold of the cross-validation, two subsets were used for training (with WSIs in the training set randomly split into training and validation sets), while the third subset served as an unseen *internal* test set. The best performing model, in terms of area under the convex hull of the receiver operating characteristic curve (AUROC) for the validation set, was chosen as the best performing trained model to obtain predictions on the held-out test set in a blinded manner.

For performance evaluation, we used AUROC, the average precision of precision-recall curve (AUPRC) and specificity values at three selected thresholds of sensitivity. All evaluation metrics were averaged for multiple folds of each experiment. We reported the mean and standard deviation of AUROC, as well as those for AUPRC. In addition, we also reported average specificity values at sensitivity values of 0·95, 0·97 and 0·99 as the benchmarks to evaluate the effectiveness and robustness of the CAIMAN tool to serve in a real-world clinical setting. We compared CAIMAN results with those obtained using IDaRS (22) with the same training and testing splits for the development and independent validation cohorts.

All performance metrics reported here were obtained from the WSI scores. To generate the slide-level label, CAIMAN employed “average” aggregation scheme, which averaged tile-level prediction scores in a slide for the abnormal classes, both non-neoplastic and neoplastic. We evaluated CAIMAN’s classification performance for non-neoplastic and neoplastic categories separately and for the abnormal category by adding non-neoplastic and neoplastic scores of each slide into a combined abnormality score. Mean and standard deviation values of all performance metrics are shown in Figure 2 for an unseen test set in a 3-fold *internal* cross-validation on the UHCW cohort (n=5054) and an independent validation on three different external cohorts, ESNE (n=148), South Warwickshire (n=257), and IMP diagnostic centre (n=1131).

**Figure 2.**
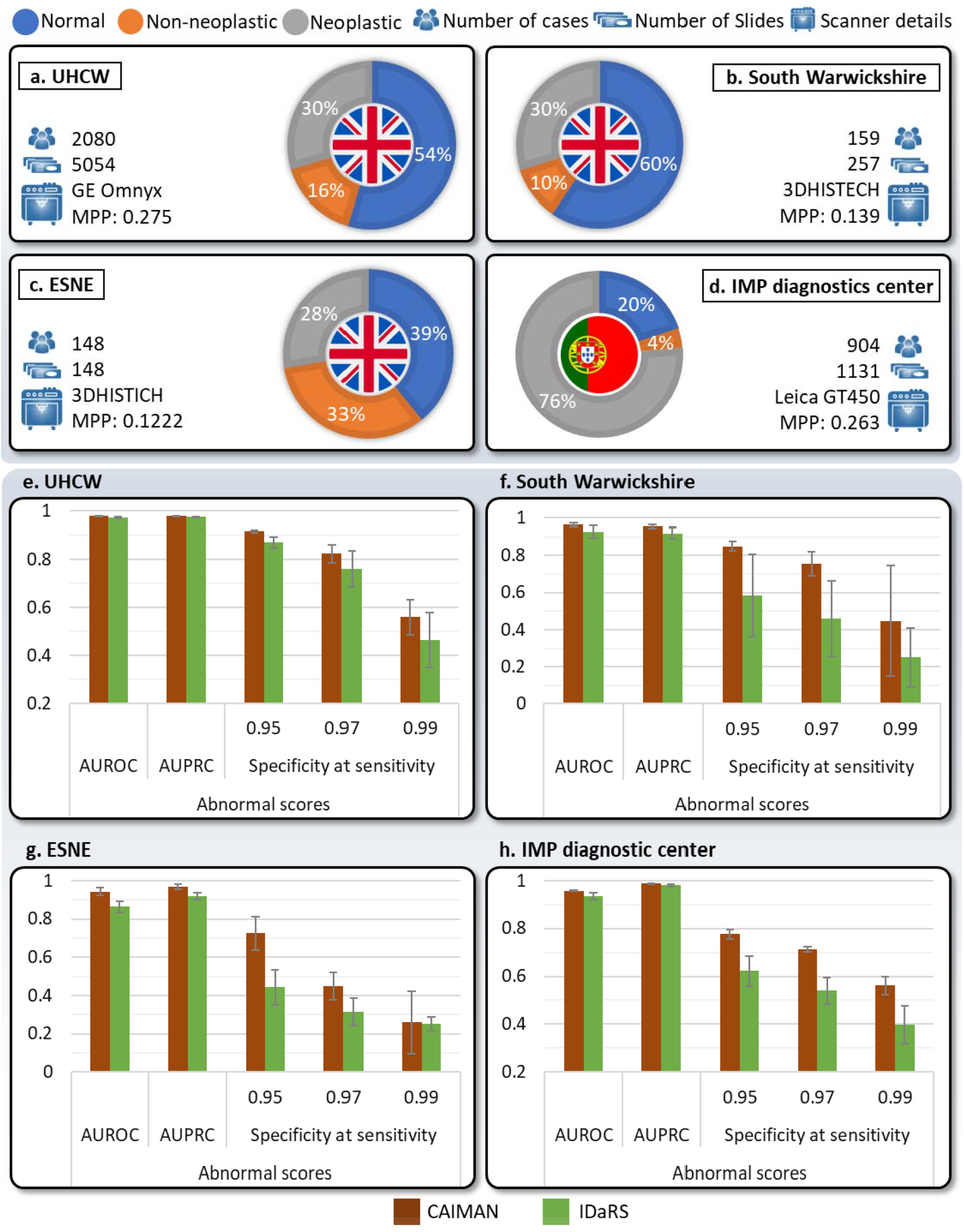
Patient cohorts used in this study (a-b.), internal cross-validation results of CAIMAN and IDaRS on UHCW (c.), and independent validation (d.) on multicentric external cohorts. Error bars show standard deviation.

Statistical significance for the difference of model prediction scores on anatomical sites of the biopsy was accessed using the Mann-Whitney U-test. The *p*-value cut-off of greater than 0·05 was used for defining not statistically significant difference. In Figure 3, we overlaid local prediction heatmaps on top of the WSIs to further analyse the relationship of diagnostic labels (normal, non-neoplastic, neoplastic) with the spatial features of the tissue microenvironment. To illustrate visual features at the tile-level corresponding to a variety of abnormality sub-conditions, we show saliency maps of strongly predicted abnormal non-neoplastic and abnormal neoplastic tiles in Figure 4. Saliency maps highlight all pixels contributing to the prediction of abnormal labels for the given image tiles.

**Figure 3.**
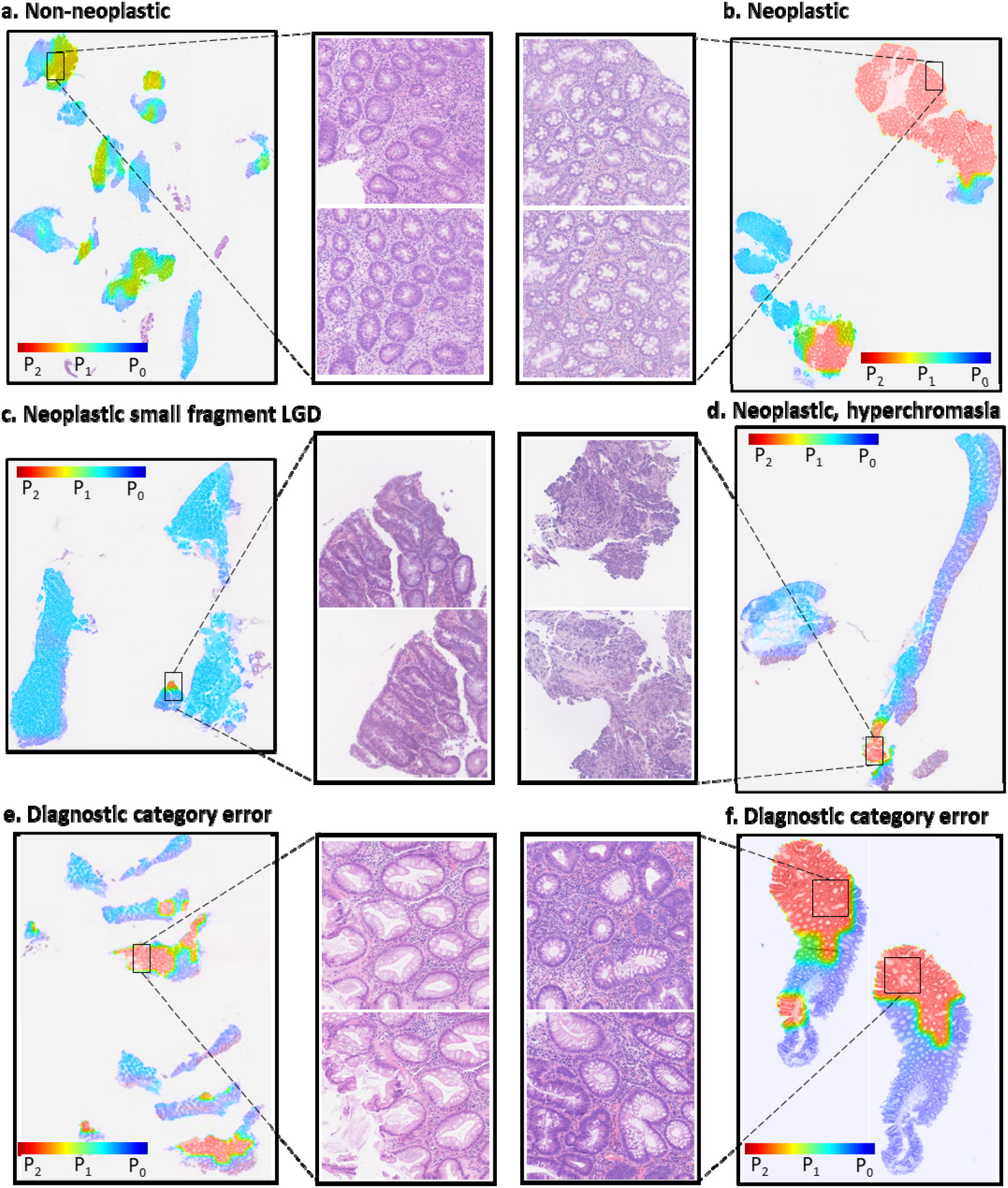
CAIMAN scores of a non-neoplastic and neoplastic slide at the tile level overlaid on original WSIs (a – d) with corresponding strongly predicted tiles, respectively; e-f. Overlay heatmaps of slides labelled with diagnostic errors but found to be predicted correctly by CAIMAN. P_2_=Neoplastic, P_1_=Non-Neoplastic and P_0_=Normal.

**Figure 4.**
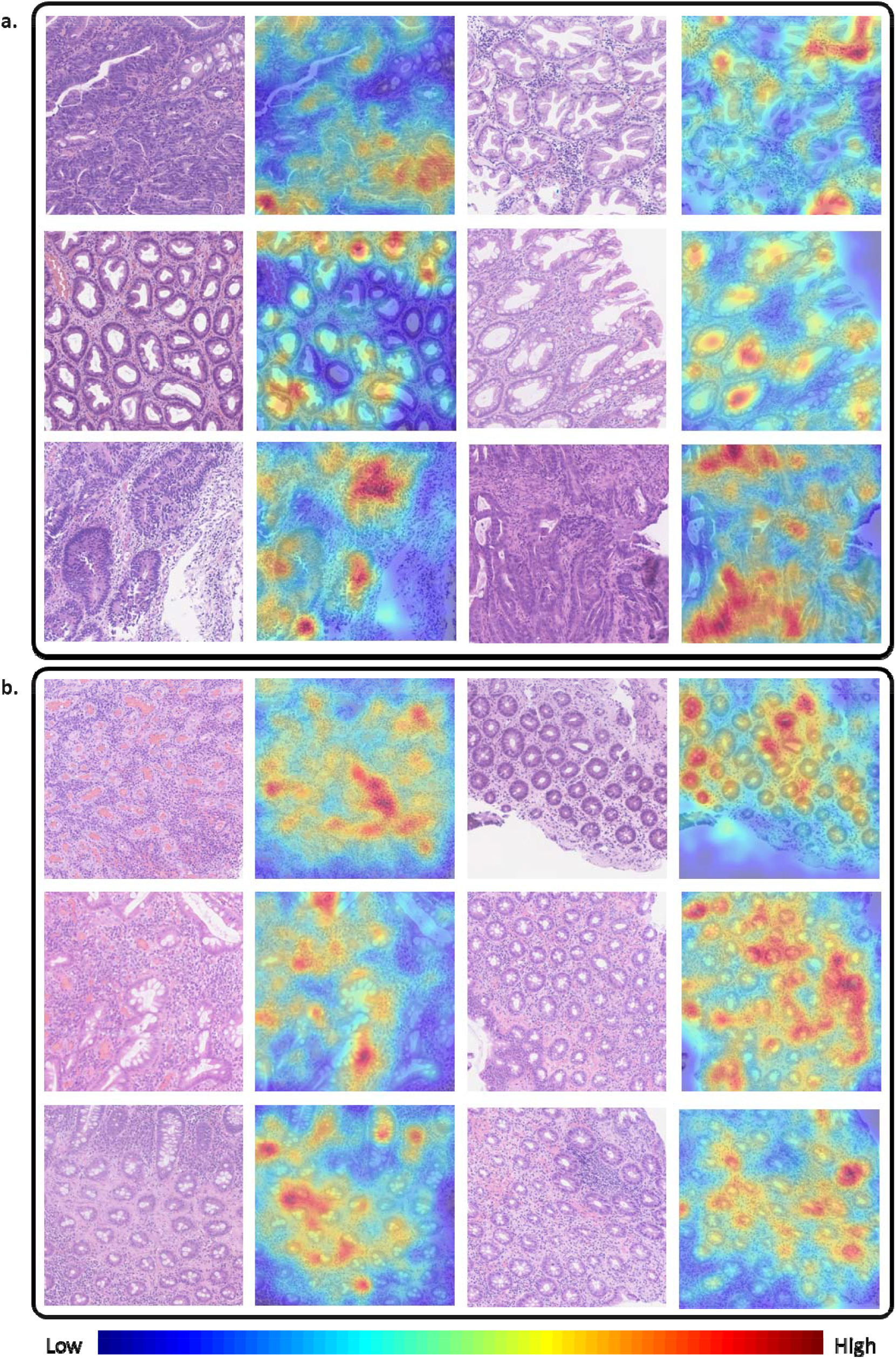
Saliency maps of example image tiles predicted accurately by CAIMAN from a. Neoplastic. and b. Non-neoplastic biopsies highlighting regions contributing towards the accurate predictions. Gaussian smoothing has been applied on the saliency maps for improved visualization.

## Results

Figure 2 provides results of CAIMAN and IDaRS for cross-validation and independent evaluation on the internal (UHCW) and external cohorts. Plots in Figure 2a-d show percentages of normal, non-neoplastic and neoplastic slides in each of the four cohorts. Results in Figure 2e-h show the performance of CAIMAN and IDaRS using AUROC, AUPRC and specificity values at sensitivity of 0·95, 0·97, and 0·99 for normal vs abnormal.

Our analysis revealed no statistically significant differences (*p*-values > 0·05) across prediction scores from the model for normal and abnormal cases based on anatomical sites of the biopsy with Mann-Whitney U-test. CAIMAN outperformed IDaRS in 3-fold internal cross-validation (Figure 2f) on the internal (UHCW) cohort in terms of all performance measures. CAIMAN also outperformed IDaRS for all independent validation cohorts (Figure 2e-g) and yielded higher specificity at selected high sensitivity cut-offs. CAIMAN produced specificities of 0·838, 0·968, 0·983 for normal vs neoplastic slides, 0·479, 0·711, and 0·80 for normal vs non-neoplastic, and 0·556, 0·825, and 0·916 for normal vs abnormal, against sensitivity values of 0·99, 0·97, and 0·95, respectively.

Figure 3a shows an overlay heatmap of tiles in a non-neoplastic slide and strongly predictive non-neoplastic tiles. Figure 3b shows a neoplastic slide and its strongly predictive neoplastic tiles. Figure 3c, d present overlay heatmaps of two interesting neoplastic slides where CAIMAN detects small neoplastic regions, one with a small fragment showing low grade dysplasia (likely carryover) and another hyperchromatic. Figure 2e, f present overlay heatmaps of two slides predicted by CAIMAN as abnormal while clinical reports suggested that they were normal.

Figure 4 shows saliency maps of strongly predicted abnormal non-neoplastic and abnormal neoplastic tiles. It is worth noting that image tiles corresponding to various abnormality sub-conditions – hyperplastic polyp, poorly differentiated adenocarcinoma, dysplasia/adenocarcinoma, high grade invasive carcinoma, as shown in Figure 4a and active inflammation, chronic inflammation, collagenous colitis, ulceration, and inflammatory bowel disease, as shown in Figure 4b – are predicted correctly by the CAIMAN model and verified by a pathologist (YWT) using the saliency maps.

## Discussion

In this study, we have proposed CAIMAN, a weakly supervised AI model for pre-screening of large bowel biopsies. The method presented in this study is aimed at assisting in the histopathological examination of large bowel biopsies in clinical practice. Our results demonstrate that AI based pre-screening offers the potential for improved efficiency as well as improved reliability. Picking a highly sensitive operating point (e.g., with sensitivity of 99%) can help greatly reduce the pathologists’ workload by automated reporting of normal cases allowing their time to be spent examining more complex cases. We found the performance difference of CAIMAN and IDaRS higher for specificity values at high sensitivity thresholds (0·95, 0·97 and 0·99) than overall AUROC and AUPRC, for both internal and external validation cohorts.

To deploy an automated colon biopsies screening system in clinical practice, a recent study (25) modelled the classification of epithelial tumours (adenocarcinoma and adenoma) with a large development cohort (n=4036) and relatively small validation cohorts (n=500, and TCGA-COAD: n=547). A more recent study (26) used gland segmentation and classification to model categorization of ‘low risk’ (benign, inflammation) and ‘high risk’ (dysplasia, malignancy) slides with a small cohort (n=294). However, these models do not cater for diagnosis of pre-cancerous lesions, non-neoplastic abnormalities and lack high sensitivity. CAIMAN can differentiate between neoplastic, non-neoplastic and normal large bowel biopsies while being able to pick up hyperplastic polyps and dysplastic lesions, thus paving the way for a reliable and more efficient pre-screening step in the routine clinical workflow with the promise of reduced pathologist workload.

CAIMAN outperforms reported accuracy figures of recently proposed supervised and semi-supervised AI tools (8,27) for CRC diagnosis of neoplastic abnormality considering multiple aspects. Wang *et al*. (8) proposed a fully supervised AI tool for cancer vs. normal prediction while Yu *et al*. (27) proposed a semi-supervised AI tool that matches the performance of supervised AI. The two methods required both slide-level and laborious tile-level annotations to achieve a sensitivity of 0·982, showing a slight improvement over an average performance of six pathologists (sensitivity = 0·975) on the same test set (8). However, it is not desirable for an AI pre-screening tool to yield high specificity at the cost of sensitivity. CAIMAN improves sensitivity up to 0·99 at the cost of slightly reduced specificity (0·84). In addition, it identifies non-neoplastic abnormalities as well. CAIMAN does not require any regional or cell level annotations for model development, thereby saving time and cost of laborious manual annotations. We have also shown that CAIMAN is able to identify a few errors in the slide-level ground truth labels. The algorithm can also pick small and sparse features of abnormality, which may be overlooked under the microscope given the vast amount of tissue content to be analysed manually.

To mitigate the effect of erroneous tile-level predictions, we implemented learning and estimating confidence at the tile level and incorporated the average slide loss in each training batch for positive slides that helped to boost the overall classification performance as well as specificity at high sensitivity threshold as compared to IDaRS (22). We did not use cellular features or richly annotated image tiles to build the deep learning model, instead relying on deep features extracted by the CAIMAN model, because of its efficient processing, automatic feature engineering and not requiring laborious detailed annotations. However, we investigated the automatically learned features by saliency and heatmap visualizations which led to identifying a few false positives at tile levels as well as errors in the ground truth labels. Inflammatory regions in both normal and neoplastic slides were also amplifying the false positives for non-neoplastic category. Supplementary Figure S2 (Appendix p.4) shows false positives, the predicted neoplastic and non-neoplastic tiles, in the normal slides. False positive rate at the tile level was mitigated with confidence learning and positive slides loss during training. On a related note, assigning a single ground-truth label to all tiles from multiple slides belonging to the same case may be interpreted as *overfitting* in classical machine learning theory. Despite the tile-level false positives, quantitative performance metrics have shown excellent predictive accuracy, robust performance and generalisation on both internal and external cohorts. This may be associated with interpolation and generalisation capabilities of over-parameterised deep learning systems leading to *benign overfitting*, as demonstrated by Bartlett *et al*. in their latest findings (28).

Limitations of this study include class imbalance in the dataset, which is often the case in medical datasets and which, it may be argued, may overrate AUROC values. To analyse the effect of class imbalance, we have also reported AUPRC measure as well. Analysis of 1% errors in identifying abnormal slides at 99% sensitivity shows that our model did not identify 22 slides of total 2294 abnormal slides, 6 out of 1500 neoplastic slides and 16 out of 794 non-neoplastic slides. During the analysis of these slides, we found 1 cancerous slide with specific diagnosis of “atypical cells-nuclear pleomorphism, hyperchromasia”, the abnormal region was detected by AI, but the average abnormality was lower because only a couple of tiles were abnormal. It further included 3 low-grade dysplasia and 2 hyperplastic slides, similar to the diagnostic errors found in the clinical reports. In the clinical reports, 5 slides of hyperplastic polyp and low-grade dysplasia was also labelled as normal. Prediction errors in non-neoplastic slides included 3 microscopic colitis, 2 inflammatory, and one each minimal focal active inflammation, mild chronic nonspecific colitis, moderate chronic and mild chronic colitis, crypt abscess, melanosis coli, isolated and tiny focus of active inflammation, patchy mild fibrosis and glandular distortion and moderate active chronic distal proctocolitis. In the normal category, we also found some normal slides with quiescent ulcerative colitis and slides with no significant inflammation.

Since the model was trained on a relatively small number of non-neoplastic slides with several sub-conditions, CAIMAN’s classification results show relatively lower performance on non-neoplastic slides. The slight drop of the generalisation performance on external validation may indicate the impact of differences because of different staining and scanners and because of varying composition of each cohort in terms of number of samples per class, which are quite different than the development cohort (UHCW) particularly in ESNE and IMP cohorts. The UHCW percentages loosely reflect the number of patients visiting UK hospitals by category.

The overall classification and generalisation performance on external cohorts can be enhanced if the amount of training data is increased with a multicentric cohort to learn from and model domain adaptation from the data of multiple different scanners and centres. We also envisage that after careful revision of the ground truth labels, both sensitivity and specificity of the model could be further improved. In future, these results merit training and validation on large and diverse multicentric cohorts, paving the way for improving the model performance and improving generalisation before its eventual clinical deployment.

While CAIMAN has demonstrated promising results, a weakly supervised model with sensitivity greater than 99% and higher level of specificity (than that demonstrated in this study) is achievable with training on larger multi-centric cohorts while ensuring that the slide-level diagnostic labels are correct and consistent across the multiple centres. Such an outcome is highly likely to assist with digital pre-screening of colorectal biopsies for multiple abnormalities in a clinical setting. As mentioned above, a digital pre-screening tool such as CAIMAN can result in time and cost efficiency, reliability and higher accuracy of routine diagnostic screening by filtering out normal large bowel biopsies from the daily workload of the pathologist and highlighting the neoplastic and non-neoplastic regions on the slide. Finally, we argue that AI-based pre-screening for large bowel biopsies is the need of the day, connecting modern digital technologies and human expertise together and consequently enabling significant decrease in cancer fatalities and reduction in under- or over-treatment as a result of timely diagnosis.

## Data Availability

Original WSIs from University Hospitals Coventry and Warwickshire NHS Trust, East Suffolk and North Essex NHS Foundation Trust and South Warwickshire NHS Foundation Trust will be made available upon completion of the PathLAKE project. Relevant information on obtaining the data from the IMP cohort can be found in the original publication.

## Acknowledgments

This paper is supported by the PathLAKE Centre of Excellence for digital pathology and artificial intelligence, which is funded from the Data to Early Diagnosis and Precision Medicine strand of the HM Government’s Industrial Strategy Challenge Fund, managed and delivered by Innovate UK on behalf of UK Research and Innovation (UKRI). The views expressed are those of the authors and not necessarily those of the PathLAKE Consortium members, the NHS, Innovate UK or UKRI.

Manuel Salto-Tellez and Jacqueline A James are Principal Investigators in PathLAKE at Queens’s University Belfast, and Clare Verrill is a Principal Investigator in PathLAKE at the University of Oxford – all were involved in generating the PathLAKE Programme, including funding.

Drs SA Sanders and N Chachlani kindly assisted in providing slides of cases from South Warwick Foundation Trust. Mr S James UHCW Tissue Bank Director assisted in scanning of slides for the external cohorts.

The authors would like to thank Johnathan Breddy, Young Park, Giorgos Hadjigeorghiou, John Euinton, Chien-Hui Liao, Thomas Leech and Valentine Stoykov for their assistance with administrative matters and IT support for software and systems utilised heavily in this project.

## Ethics Approval and Consent to Participate

This study was conducted under Health Research Authority National Research Ethics approval 15/NW/0843; IRAS 189 095 and the Pathology image data Lake for Analytics, Knowledge and Education (PathLAKE) research ethics committee approval (REC reference 19/SC/0363, IRAS project ID 257 932, South Central - Oxford C Research Ethics Committee). Data collection and usage of the IMP Diagnostics dataset was performed in accordance with national legal and ethical standards applicable to this cohort (21).

## Contributors

MB, DS and NR designed the study with support from all co-authors. MB and NR developed the methods. MB wrote the code and carried out all experiments. YWT, KG, MA, EH, AR, AA, EH, KD, HS, MN, KH, KB and DS provided diagnostic annotations of colonic biopsy slides. YWT, MB, FM, KG, and MA and performed analysis and interpretation of the results. MJ and MB made visualisations. FM, SEAR, AB, SG, MJ, NW and WL provided technical and material support. MB, YWT, FM, DS and NR were all involved in drafting of the paper. DS and NR jointly supervised the study and performed analysis and interpretation of the results. MB and NR had access to all the data while other authors could gain access to the data as and when needed. All authors agreed to proceed with submission of the manuscript.

## Data availability statement

Original WSIs from University Hospitals Coventry and Warwickshire NHS Trust, East Suffolk and North Essex NHS Foundation Trust and South Warwickshire NHS Foundation Trust will be made available upon completion of the PathLAKE project. Relevant information on obtaining the data from the IMP cohort can be found in the original publication (21). The scores generated by our algorithm are provided in appendix. Source code can be made available, subject to intellectual property constraints, by contacting the first or last authors.

## Footnotes

2 https://www.bowelcanceruk.org.uk/about-bowel-cancer/bowel-cancer/

